# Online Peer Support Use Among Informal Caregivers of People Living With Alzheimer Disease or Related Dementias: National Cross-Sectional Web-Based Survey

**DOI:** 10.64898/2026.09.02.26362079

**Authors:** Congning Ni, Lantian Xia, Lijun Song, Qingxia Chen, Sean S. Huang, Bradley Malin, Zhijun Yin

## Abstract

**Background:** Informal caregivers of persons living with Alzheimer’s disease or related dementias (ADRD) increasingly seek support in online environments. Prior work suggests that perceived value may shape online support use, yet practical and trust-related barriers may still impede uptake even when beliefs are favorable.

**Objective:** To characterize patterns of online peer support use among informal ADRD caregivers and identify and analyze the predisposing, enabling, and need factors under the Andersen and Newman Framework of Health Services Utilization (ANFHSU) framework that are associated with use and nonuse behaviors.

**Methods:** We conducted a cross-sectional, web-based survey that recruited a national, nonprobability sample of US adults through Qualtrics online panels from July 15 to September 16, 2023. Online peer support use was defined as searching for information or discussing caregiving topics in online communities during the past 3 months. Guided by ANFHSU, we measured predisposing characteristics, enabling resources, and need-related factors, along with additional technology-related preference measures, including willingness to use an experience-search tool and a caregiving-education tool. We summarized patterns in platform ecosystem and engagement modes and fitted 5 nested multivariable logistic regression models to identify factors associated with recent online peer support use. Among non-users, we evaluated multiple thresholds to identify a high-belief subgroup, summarized reported barriers, and modeled intention to use online peer support in the next 3 months.

**Results:** Of 18,245 invited panel participants, 12,072 completed the survey (participation rate, 66.2%). After eligibility screening and data-quality exclusions, 1,113 unpaid informal ADRD caregivers comprised the analytic sample. Overall, 740/1,113 (66.5%) caregivers reported online peer support use in the past 3 months. Use was distributed across a fragmented platform ecosystem, with a long tail of infrequently named sources. More than half of users (402/740, 54.3%) primarily read rather than wrote posts, indicating substantial passive participation. In the fully adjusted model, higher belief in the value of online peer support was associated with greater odds of use (OR 1.04, 95% CI 1.01-1.08), along with higher eHealth literacy (OR 1.06, 95% CI 1.02-1.10), greater willingness to use an experience-search tool (OR 1.68, 95% CI 1.39-2.04), and higher caregiving stress (OR 1.03, 95% CI 1.01-1.05). Among non-users (373/1,113, 33.5%), 108/373 (28.9%) met the retained high-belief threshold despite reporting no recent use, indicating a belief-behavior gap. Among non-users, the most commonly reported barriers were sufficient offline support (118/373, 31.6%), lack of time (102/373, 27.3%), dislike of online communities (79/373, 21.2%), limited opportunity to join despite awareness (74/373, 19.8%), and security or trust concerns (67/373, 18.0%). Among non-users, 161/373 (43.2%) reported intending to use online peer support in the next 3 months.

**Conclusions:** Our findings suggest that uptake of online peer support reflects perceived value, digital readiness, and caregiving-related need rather than need alone. A meaningful belief-behavior gap indicates that favorable beliefs do not ensure use when time, trust, and access barriers persist. These findings suggest that efforts to support ADRD caregivers should focus on reducing practical and trust-related barriers to online peer support and should recognize that engagement often occurs through reading rather than posting.

## 1 Introduction

Alzheimer’s disease or related dementias (ADRD) impose profound challenges not only on those diagnosed but also on informal caregivers such as family members and friends who provide most of the daily care [1]. Nearly 12 million Americans are currently providing unpaid care for a person living with ADRD, contributing an estimated 18 billion hours of care in 2023 [2]. This caregiving role often spans several years and is associated with substantial emotional stress, physical strain, and financial hardships [3,4]. As the population ages and ADRD prevalence rises, supporting caregiver well-being has become a public health priority [5]. Traditional in-person support services (such as support groups, respite care, and educational programs) exist [6]. However, many caregivers face numerous obstacles in accessing in-person support, including conflicts between caregiving and work responsibilities, geographical distance from service providers, and disruptions to service provision during and after the COVID-19 pandemic [7]. Caregiver burden can also accumulate as emotional distress, grief, and depressive symptoms, further increasing the need for flexible and timely sources of support [8].

Online peer support communities range from specialized forums (e.g., ALZConnected and the Alzheimer’s Society Dementia Support Forum) to broader social platforms and open discussion spaces (e.g., Facebook and Reddit). These online communities have become important venues for caregivers to seek information and emotional support from others facing similar situations [9]. Unlike professional consultation services, online peer support allows caregivers to exchange practical caregiving tips, express frustration, and receive encouragement at any time, without the constraints of location or scheduled appointments [10,11]. Prior research on web-based caregiver support suggests that participation in online communities may reduce feelings of isolation, increase knowledge about managing dementia-related behaviors, provide empathetic understanding that may be missing from offline networks [12,13], and improve coping skills and confidence [14]. More broadly, peer-support models have shown benefit in other health contexts, e.g., alcohol use disorder, suggesting that support grounded in shared lived experience can complement formal services [15].

Despite the availability and benefits of online support platforms, many ADRD caregivers do not regularly use online peer support [16]. Prior research indicates a substantial digital divide in this domain: caregivers who are older, have lower digital skills, or are from underserved communities are less likely to use online resources [17]; and current online community users tend to be skewed toward middle-aged female caregivers with higher education [14]. Reported barriers include limited awareness of online support options, low trust in internet information, limited computer or smartphone skills, and simply not having enough time or energy to go online after caregiving responsibilities [16–18].

Meanwhile, existing studies have often treated online peer support use as a relatively simple yes-or-no behavior [21] and have paid less attention to how caregivers engage once they arrive, including whether they primarily read discussions or actively post and respond. Moreover, existing studies have examined these barriers descriptively [14] or within specific subgroups [22], without a systematic, theory-guided assessment of how individual beliefs, enabling resources, and caregiving-related needs jointly shape online peer support use [23]. To address this gap, we previously conducted a survey study guided by the Andersen and Newman Framework of Health Services Utilization (ANFHSU) [24], which conceptualizes service use as a function of predisposing characteristics, enabling resources, and perceived need. We found that caregivers’ belief in the value of online peer support was the strongest predictor of use, even though most caregivers who did not engage online still reported positive beliefs about its potential usefulness [25]. However, our prior survey had several limitations that constrained broader inference. The sample size was modest, recruitment relied primarily on the Alzheimer’s Association website and ALZConnected, and the analysis did not examine how digital capability, trust-related concerns, and caregiving context interact at scale. As a result, it remained unclear whether the previously observed belief-behavior gap would persist in a larger sample, how recent online peer support use would vary across platform ecosystems and participation modes, and which enabling or need-related factors would remain independently associated with use after multivariable adjustment.

In this study, we extended the prior survey into a nationwide survey of 1,113 informal ADRD caregivers. In addition to core predisposing, enabling, and need factors assessed previously, we incorporated measures of dementia-related knowledge, eHealth literacy, and technology-related preference measures, including willingness to use an experience-search tool and a caregiving-education tool to better capture caregivers’ capacity to locate, evaluate, and use digital support resources in a contemporary information environment. We characterize where and how caregivers engage in online peer support, including fragmented platform use and patterns of passive versus active participation. We further identify ANFHSU-aligned factors associated with recent online peer support use, with particular attention to caregiver beliefs, digital readiness, and caregiving burden. Finally, we examine nonuse among caregivers who nevertheless perceive online peer support as valuable, focusing on practical, trust-related, and access-related barriers that may sustain a belief-behavior gap.

## 2 Methods

### 2.1 Study Design and Ethics

We conducted a cross-sectional, web-based survey to examine informal dementia caregivers’ perceptions of, access to, and engagement with online peer support. The survey was conducted through Qualtrics and administered as a self-completed questionnaire accessible via computer or mobile device. All responses were collected electronically and anonymously. This study was reviewed and deemed non-human subjects research by the Vanderbilt University Medical Center Institutional Review Board (IRB #221732).

### 2.2 Participants, Recruitment, and Data Collection

Eligible participants were adults who met all the following criteria: (1) reported knowledge of Alzheimer’s disease or related dementias, (2) reported that they were currently providing or had recently provided unpaid care to a person living with Alzheimer’s disease or related dementias, and (3) reported receiving no financial compensation for caregiving. In the survey, caregiving was defined as assisting with activities of daily living, including eating, bathing, transportation, and attending medical visits. Respondents who did not meet eligibility criteria were excluded through programmed screening and skip logic embedded in the questionnaire.

We fielded the survey through Qualtrics online panels from July 15 to September 16, 2023, recruiting a national, nonprobability sample of US adults. Potential respondents accessed the web-based survey through Qualtrics’ panel infrastructure, completed eligibility screening, and proceeded to the main questionnaire only if they met study criteria. Survey responses were collected electronically and stored in a secure database. Each response was associated with a study-specific identifier, and no direct personal identifiers were retained in the analytic dataset. Prior to analysis, we applied prespecified data-quality screening criteria, including exclusions for substantial noncompletion, implausibly short completion times suggestive of non-engaged responding, and failed attention checks.

### 2.3 Survey Instrument Development

Social support is associated with better coping and health outcomes, particularly under sustained stress and complex care demands[26]. This study was guided by ANFHSU, which frames service use as a function of predisposing characteristics, enabling resources/capabilities, and care-related need. We conceptualized online peer support as a health-related support resource and designed the questionnaire to operationalize these domains. To reduce ambiguity in respondents’ interpretations of online engagement, the survey defined “online community” as an online space where people with similar experiences or knowledge discuss with one another (e.g., social platforms, forums, and condition-specific communities).

#### 2.3.1 Predisposing factors

Predisposing factors capture caregivers’ background characteristics, caregiving context, and attitudes that may shape the propensity to seek support resources. Specifically, we measured:

- **Caregiver sociodemographic and household context.** Respondents reported age, sex, race, ethnicity, living area, education, employment status, household income, marital status, number of children under care (where applicable), and household size. These variables describe structural and contextual characteristics linked to health information-seeking and digital resource use.
- **Caregiving relationship and proximity**. Respondents reported their relationship to the care recipient (e.g., spouse/partner, adult child, other relative, friend/neighbor) and whether they lived in the same neighborhood as the care recipient. These items capture role expectations and proximity-related constraints that may influence support-seeking preferences [27].
- **Care-recipient characteristics and dementia stage.** Respondents reported basic care-recipient characteristics (e.g., age, sex, marital status) and caregiver-perceived dementia stage (e.g., mild/moderate/severe). Dementia stage provides context for anticipated care complexity and support needs.
- **Self-rated health status.** Respondents indicated agreement with “I feel healthy and do not have any major diseases that affect my daily life,” using an ordinal agreement scale. This item captures baseline caregiver health as a potential correlate of capacity and engagement in support-seeking.
- **Beliefs about the value of online peer support.** Attitudes toward online peer support were assessed using a 6-item belief battery asking how likely reading online discussions from, or writing posts to communicate with, other caregivers in online communities would help respondents find needed resources, increase understanding of the disease and patient, improve caregiving skills, increase caregiving confidence, reduce caregiving stress, and reduce loneliness. Each item used a 7-point response scale ranging from extremely unlikely to extremely likely, and items were combined into a composite belief score, with higher scores indicating more favorable beliefs about online peer support [28].

#### 2.3.2 Enabling factors

Enabling factors capture access conditions and digital capability that facilitate (or impede) engagement with online peer support. In this survey, we measured:

- **Perceived internet adequacy.** Respondents rated agreement with “My current internet speed is fast enough for me to search for information or connect with other people online,” using an ordinal agreement scale [29].
- **eHealth Literacy Scale (eHEALS) [30].** Respondents completed a multi-item eHealth literacy battery assessing perceived ability to locate, evaluate, and apply online health information (e.g., knowing where/how to find helpful resources, distinguishing high- vs low-quality information, confidence using online information for decisions). Items used an ordinal agreement scale.

#### 2.3.3 Need factors (care demands and caregiver strain)

Need factors capture caregiving demands, strain, and informational needs that may motivate additional support-seeking. In this survey, we measured:

- **Caregiving duration and frequency.** Respondents reported how long they had been caring for the PLWD and how frequently they provided care each week. These variables were analyzed separately to capture different aspects of caregiving demand.
- **Caregiver burden/stress.** Caregiver strain was assessed using a 12-item caregiver burden battery with frequency-based response options (e.g., role strain, time constraints, emotional strain, impacts on social life and health, uncertainty, self-criticism). A total stress/burden score was computed by summing item responses according to the Zarit Burden Interview (12-item) scale [31].
- **Caregiving competence/confidence**. Caregiver competence was assessed using a brief multi-item battery capturing perceived confidence and competence in managing caregiving challenges. A composite competence score was computed by summing item responses [32].
- **Dementia-related knowledge.** Dementia knowledge was assessed using a structured knowledge battery consisting of statements about Alzheimer’s disease/dementia and caregiving. Responses were scored using the survey’s prespecified scoring rule to generate a composite knowledge score, with higher scores indicating greater dementia-related knowledge.
- **Caregiving challenges.** Respondents selected major caregiving challenges using a “select all that apply” checklist spanning practical, emotional, clinical, communication, safety, family, legal or financial, and role-balance domains. These items were included to characterize the types of caregiving demands respondents faced and to contextualize perceived need for additional support [33].
- **Satisfaction with support from multiple sources.** Respondents rated their satisfaction with support received from a structured set of sources, including family and friends, local and online dementia caregivers, clinicians and allied health professionals, community and nonprofit resources, legal services, and self-learning on caregiving [26]. Responses were recorded in a matrix with options ranging from quite unsatisfied to quite satisfied, plus a not-applicable option. For analysis, satisfaction with peer support in online forums was retained as a distinct measure, and satisfaction with non-peer support sources was summarized separately to capture the perceived adequacy of existing offline and formal support.

#### 2.3.4 Trust, Misinformation Concerns, and Tool-Related Preferences

To inform intervention-oriented implications related to trust and information credibility, the survey assessed caregivers’ willingness to use two types of digital support tools.

- **Willingness to use experience-search tools.** Respondents rated their likelihood of using a tool that could help caregivers efficiently locate relevant online discussions or connect with caregivers who had similar experiences [34].
- **Willingness to use caregiving-education tools.** Respondents rated their likelihood of using a tool that could provide clear, correct explanations for common misunderstandings related to ADRD and caregiving [35].

Both items used ordinal likelihood scales and were analyzed as willingness measures rather than indicators of current app use or existing information-seeking behavior.

### 2.4 Statistical Analysis

We conducted analyses to address three primary hypotheses regarding the use of online peer support among dementia caregivers.

**H1: Caregiver characteristics, beliefs, and contextual factors differ between users and non-users of online peer support.** We described caregiver and care recipient characteristics, online peer support use patterns, and key study measures of recent online peer support use over the past 3 months. Categorical variables were summarized using frequencies and percentages, and continuous variables were summarized using means and standard deviations. Group differences between recent users and non-users were examined using chi-square tests for categorical variables, with Fisher’s exact tests used for sparse 2×2 tables when needed. Continuous variables were compared using independent-samples *t* tests when group distributions approximated normality, and Mann-Whitney *U* tests otherwise. Effect sizes were summarized using Cramer’s *V* for categorical comparisons, Cohen’s *d* for *t*-test comparisons, and rank-based effect sizes for Mann-Whitney comparisons.

**H2: ANFHSU-aligned factors are associated with online peer support use, and particularly belief in the value of online peer support is independently associated with use.** To identify factors associated with online peer support use in the past three months, we fitted ANFHSU-guided multivariable logistic regression models in a blockwise sequence. Specifically, we estimated five nested models: a predisposing-only model excluding belief measures (M1), then sequentially added belief scores regarding online peer support (M2), enabling factors including perceived internet adequacy and eHealth literacy (M3), need-related factors including caregiving burden and competence (M4), and additional knowledge-, technology-, and support-related variables (M5), including willingness to use an experience-search tool and willingness to use a caregiving-education tool. Before modeling, variables were coerced to numeric where appropriate, and sparse categorical levels were collapsed to improve model stability. Multicollinearity among numeric predictors was assessed using variance inflation factors. Standard logistic regression was used as the default estimation approach, and regularized logistic regression was used when numerical instability or convergence problems occurred. Model performance was evaluated using Akaike information criterion (AIC), McFadden’s pseudo-*R*², and ROC AUC.

**H3: Among caregivers who did not report online peer support use in the past 3 months, those with higher belief in the value of online peer support differ from those with lower belief, consistent with a belief-behavior gap subgroup.** To examine the belief-behavior gap, we restricted analyses to caregivers who did not report online peer support use in the past three months. We evaluated multiple thresholding approaches to define a high-belief subgroup, including percentile-based and fixed-score thresholds, and retained the threshold used for detailed reporting in the current analysis. High-belief non-users were then compared with the remaining non-users who did not meet the high-belief threshold on continuous variables of interest, including eHealth literacy, caregiving stress, caregiving competence, ADRD knowledge, satisfaction with existing support, and willingness to use digital support tools, using independent-samples *t* tests and Cohen’s *d*. Reported reasons for nonuse were summarized descriptively. Among non-users, we additionally modeled intention to use online peer support in the next three months as a secondary binary outcome using a logistic regression framework aligned with the same ANFHSU variable blocks.

Internal consistency was assessed for composite scales for which item-level data were available in the analytic dataset, including the belief scale and the support-satisfaction scale, using Cronbach’s alpha. Missing data were handled using available-case analysis for descriptive and univariable comparisons and complete-case estimation within each regression model; therefore, analytic sample sizes vary across models and are reported with results. All analyses used a two-sided α of 0.05.

## 3 Results

### 3.1 Characteristics of Caregivers and Care Recipients

Of 18,245 invited panel participants, 12,072 completed the survey (participation rate 66.2%). After eligibility screening and data-quality exclusions, 1,113 unpaid informal ADRD caregivers comprised the analytic sample. Overall, 740/1,113 (66.5%) caregivers reported using online peer support in the past 3 months, and 373/1,113 (33.5%) reported no recent use. To address H1, we compared caregiver and care-recipient characteristics between users and non-users of online peer support.

Caregiver characteristics are summarized in Table 1. Specifically, caregivers had a mean age of 46.4 ± 16.1 years and were predominantly female (777/1,113, 69.8%), White (869/1,113, 78.1%), and not Hispanic/Latino (962/1,113, 86.4%). Living area was distributed across small-town/rural (481/1,113, 43.2%), urban core (279/1,113, 25.1%), suburban (269/1,113, 24.2%), and large rural (84/1,113, 7.5%). Educational attainment spanned multiple levels, with the largest groups reporting some college/2-year degree (374/1,113, 33.6%), high school/GED (278/1,113, 25.0%), or a 4-year college degree (274/1,113, 24.6%). Approximately half were employed full-time (564/1,113, 50.7%), and household income covered a wide range, including $25,000-$49,999 (314/1,113, 28.2%) and <$25,000 (253/1,113, 22.7%). Caregivers reported 1.66 ± 0.71 children under care and a mean household size of 3.33 ± 1.78.

**Table 1.**
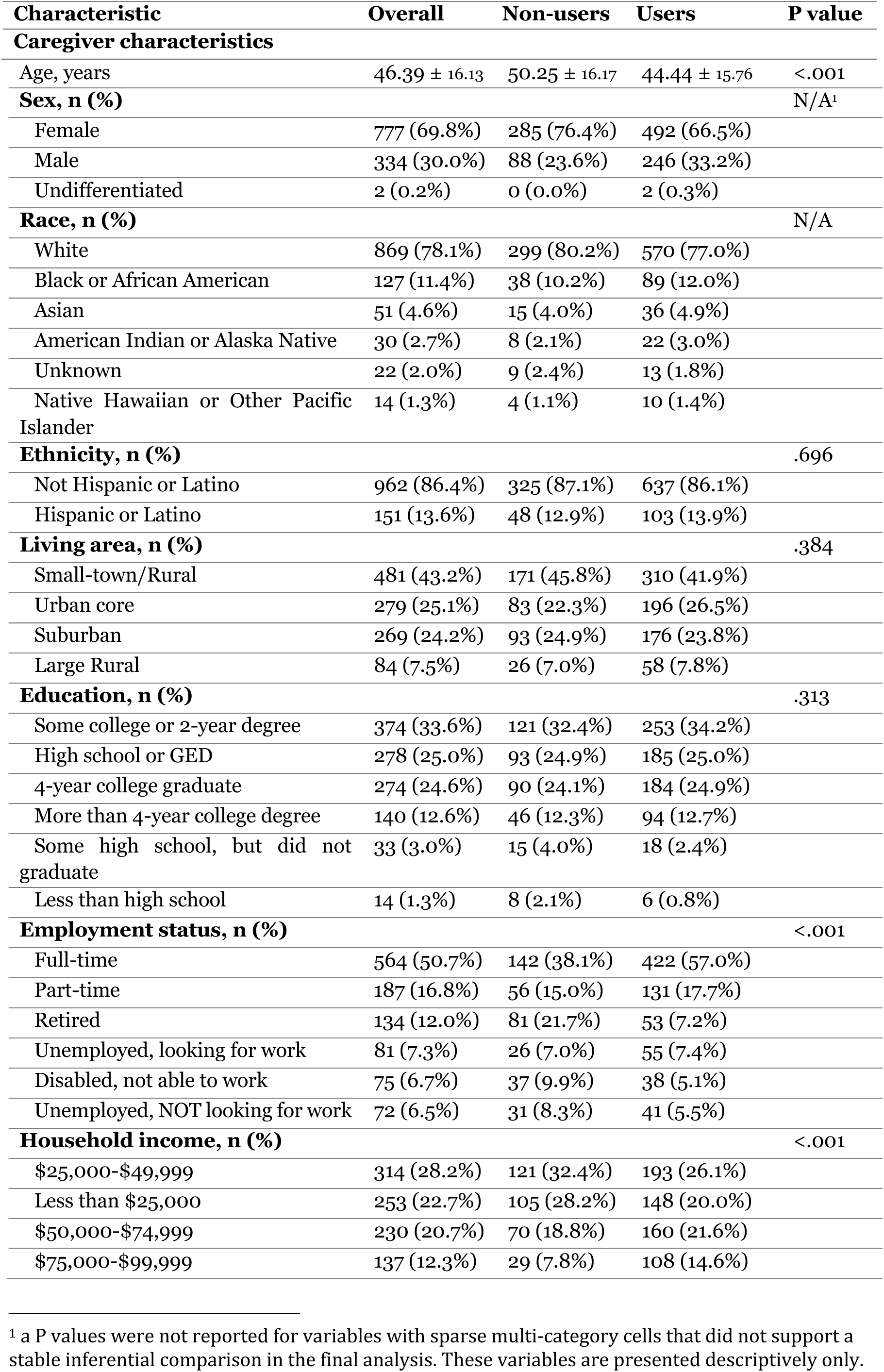
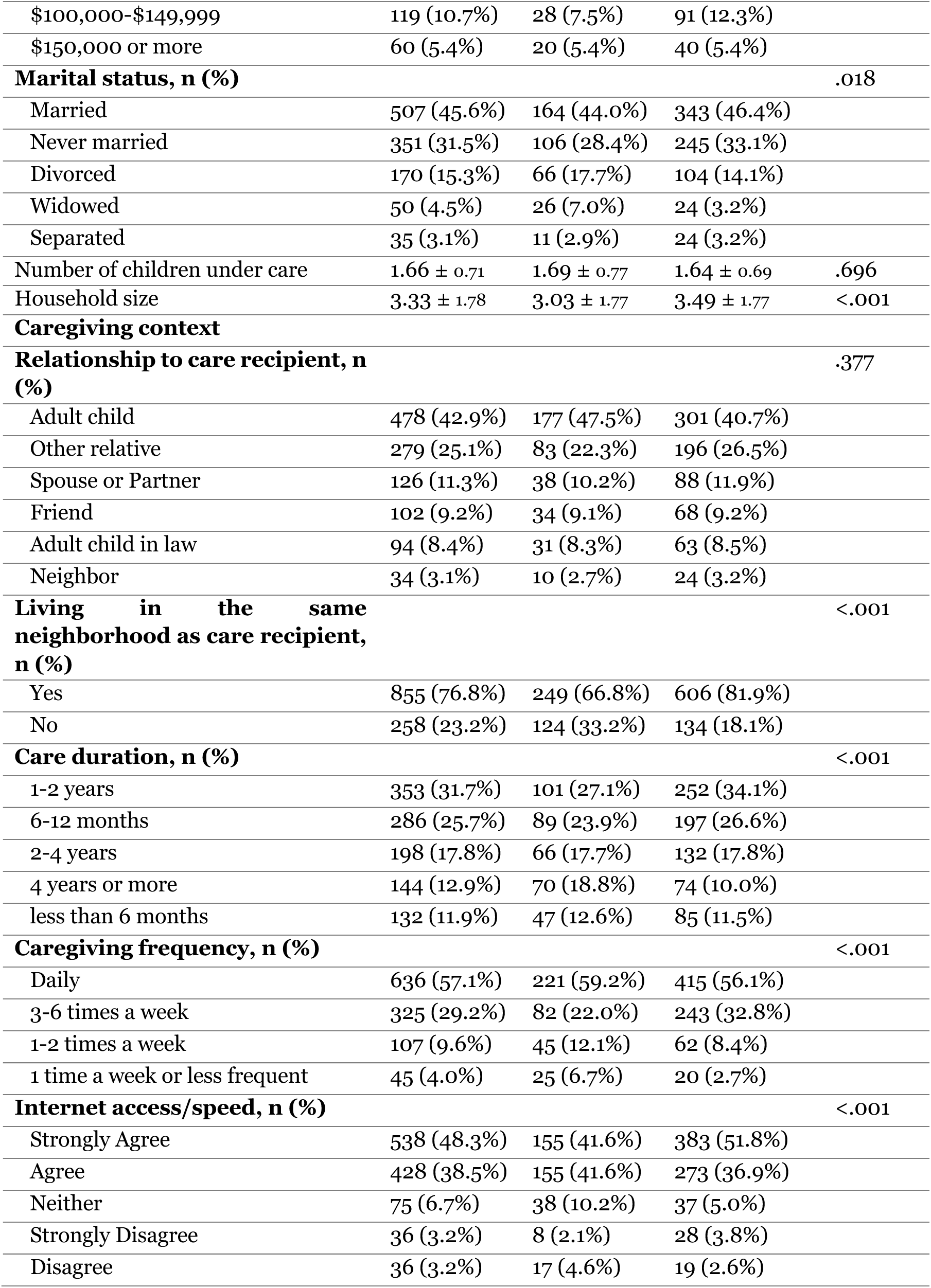
Caregiver characteristics of the analytic sample (N = 1,113) by online peer support use in the past 3 months. Values are reported as mean ± SD or n (%). P values reflect unadjusted comparisons between non-users (n = 373) and users (n = 740).

Caregiving context measures indicated that the most common relationships to the care recipient were adult children (478/1,113, 42.9%) and other relatives (279/1,113, 25.1%). Most caregivers lived in the same neighborhood as the care recipient (855/1,113, 76.8%). Caregiving duration spanned a wide range, including less than 6 months (132/1,113, 11.9%), 6-12 months (286/1,113, 25.7%), 1-2 years (353/1,113, 31.7%), 2-4 years (198/1,113, 17.8%), and 4 years or more (144/1,113, 12.9%). Caregiving intensity was high, with daily caregiving (636/1,113, 57.1%) being most frequent, followed by 3-6 times/week (325/1,113, 29.2%). For perceived internet access/speed, most respondents selected either strongly agree (538/1,113, 48.3%) or agree (428/1,113, 38.5%).

Care-recipient (PLWD) characteristics are summarized in Table 2. PLWDs had a mean age of 72.8 ± 15.2 years; 60.7% were female (676/1,113) and the most common marital status was widowed (521/1,113, 46.8%). Dementia stage was most frequently reported as middle-stage (moderate) (617/1,113, 55.4%), followed by early-stage (mild) (251/1,113, 22.6%) and late-stage (severe) (245/1,113, 22.0%).

**Table 2.** Care-recipient (PLWD) characteristics of the analytic sample (N = 1,113) by online peer support use in the past 3 months. Values are reported as mean ± SD or n (%). P values reflect unadjusted comparisons between non-users (n = 373) and users (n = 740)

| **Characteristic** | **Overall** | **Non-users** | **Users** | **P value** |
| --- | --- | --- | --- | --- |
| Care recipient age, years | 72.80 ± 15.22 | 75.37 ± 14.38 | 71.50 ± 15.48 | <.001 |
| **Care recipient sex, n (%)** |  |  |  | N/A |
| Female | 676 (60.7%) | 226 (60.6%) | 450 (60.8%) |  |
| Male | 434 (39.0%) | 146 (39.1%) | 288 (38.9%) |  |
| Undifferentiated | 3 (0.3%) | 1 (0.3%) | 2 (0.3%) |  |
| **Care recipient marital status, n (%)** |  |  |  | .535 |
| Widowed | 521 (46.8%) | 180 (48.3%) | 341 (46.1%) |  |
| Married | 341 (30.6%) | 120 (32.2%) | 221 (29.9%) |  |
| Divorced | 140 (12.6%) | 39 (10.5%) | 101 (13.6%) |  |
| Never married | 77 (6.9%) | 24 (6.4%) | 53 (7.2%) |  |
| Separated | 34 (3.1%) | 10 (2.7%) | 24 (3.2%) |  |
| **Dementia stage, n (%)** |  |  |  | <.001 |
| Middle-stage (moderate) | 617 (55.4%) | 170 (45.6%) | 447 (60.4%) |  |
| Early-stage (mild) | 251 (22.6%) | 98 (26.3%) | 153 (20.7%) |  |
| Late-stage (severe) | 245 (22.0%) | 105 (28.2%) | 140 (18.9%) |  |

When stratified by online peer support use status (Tables 1, 2), several differences were observed. Compared with non-users, users were younger (44.4 ± 15.8 vs 50.3 ± 16.2 years; P<.001) and lived in larger households (3.5 ± 1.8 vs 3.0 ± 1.8; P<.001), while the number of children under care did not differ (P = .696). Ethnicity distributions did not differ (P = .696), and differences by living area and education were not significant (P = .384 and P = .313, respectively). Employment and income patterns differed significantly: users were more likely to be employed full-time (57.0% vs 38.1%) and less likely to be retired (7.2% vs 21.7%; P<.001), and they were less concentrated in the lowest income category (<$25,000: 20.0% vs 28.2%) with higher representation in middle-to-upper income brackets (e.g., $75,000-$99,999: 14.6% vs 7.8%; overall P<.001). Marital status also differed modestly (P = .018). In caregiving context, users were more likely to live in the same neighborhood as the care recipient (81.9% vs 66.8%; P<.001), and they were less likely to report very long caregiving durations (≥4 years: 10.0% vs 18.8%; P<.001). Caregiving frequency also differed (P<.001), with users more often reporting caregiving 3-6 times per week (32.8% vs 22.0%) and less often reporting low-frequency caregiving (e.g., 1 time/week or less: 2.7% vs 6.7%). Users also more frequently reported strong internet access/speed (strongly agree: 51.8% vs 41.6%; overall P<.001).

Care recipients of users were, on average, younger than those of non-users (71.5 ± 15.5 vs 75.4 ± 14.4 years; P<.001) (Table 2). Dementia stage distributions also differed (P<.001): users more often cared for PLWDs in the middle stage (60.4% vs 45.6%) and less often in the late stage (18.9% vs 28.2%), while care-recipient marital status was similar between groups (P = .535).

### 3.2 Online Peer Support Use, Platforms, and Engagement

Among the 1,113 caregivers included in the analytic sample, 740 (66.5%) reported using online peer support in the past three months, whereas 373 (33.5%) reported no recent use. As part of H1, we summarized recent online peer support use, platform ecosystem patterns, visit frequency, and engagement mode.

**Platform Ecosystem**. Caregivers who reported online peer support use engaged across a diverse set of online venues (Figure 1). Across coded responses, we identified 14 platform categories. Among users with codable platform entries, most reported a single platform category (mean 1.24 platform categories per user), whereas 141/740 (19.1%) reported using multiple platform categories (maximum 5 platform categories reported by a single respondent). When summarized by unique users, the largest share of caregivers reported platforms coded as “Other” (460/740, 62.2%), reflecting a long tail of less frequently named or heterogeneous sources. Among specifically named platforms, Facebook was most commonly reported (129/740, 17.4%), followed by Reddit (75/740, 10.1%) and web forums (52/740, 7.0%). Smaller proportions reported using Google (46/740, 6.2%), medical sites (32/740, 4.3%), and ALZConnected (24/740, 3.2%), with each remaining category reported by fewer than 3% of users.

**Figure 1.**
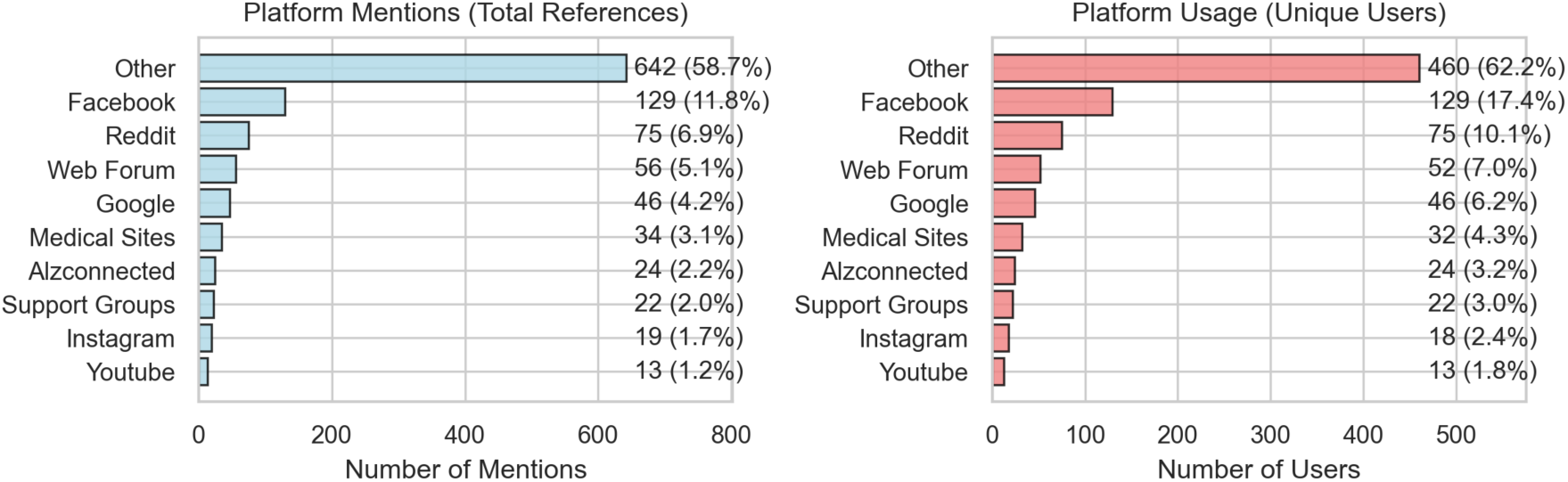
Platform ecosystem of online peer support use among caregivers reporting use in the past three months (n=740). Bars summarize total platform references (left) and the number of unique users mentioning each platform category (right).

**Visit Frequency.** Among caregivers who used online peer support, reported visit frequency varied. The largest proportion of users (310/740, 41.9%) reported visiting at least once per week. Additional users reported visiting at least once per month (157/740, 21.2%) or daily (133/740, 18.0%). A further 140/740 (18.9%) reported visiting only when a specific caregiving question or concern arose.

**Engagement Mode.** Caregivers differed in how actively they participated within online peer support spaces. More than half of users (402/740, 54.3%) reported primarily reading discussions between other caregivers without posting. Another 201/740 (27.2%) reported both reading and writing posts or comments, whereas 137/740 (18.5%) reported primarily communicating by writing posts or comments.

**Intention to Revisit Online Peer Support.** Among caregivers who reported recent online peer support use, 667/740 (90.1%) reported that they intended to visit online communities again in the next three months. This suggests that online peer support may continue to function as an ongoing support resource for many current users.

### 3.3 Key Study Measures by Online Peer Support Use

To further address H1, we compared key continuous study measures between caregivers who did and did not report online peer support use in the past three months (Table 3). Compared with non-users, users reported higher belief in the value of online peer support, higher eHealth literacy, higher caregiver stress, higher caregiving competence, and higher satisfaction with non-peer support sources. By contrast, non-users had slightly higher ADRD knowledge scores than users.

**Table 3.** Key study measures by online peer support use in the past 3 months. Values are reported as mean (SD). P values reflect unadjusted comparisons between non-users (n = 373) and users (n = 740); all comparisons used Mann-Whitney U tests. Effect sizes are rank-based.

| Measure | Overall | Non-users | Users | P value | Effect size |
| --- | --- | --- | --- | --- | --- |
| Belief in the value of online peer support | 26.94<br>(6.38) | 24.56<br>(6.54) | 28.14<br>(5.94) | <.001 | 0.343 |
| eHealth literacy | 27.89<br>(5.49) | 26.20<br>(5.55) | 28.74<br>(5.27) | <.001 | 0.275 |
| Caregiver stress | 17.85<br>(9.66) | 16.75<br>(9.68) | 18.40<br>(9.61) | .005 | 0.103 |
| Caregiving competence | 8.93 (2.37) | 8.65 (2.51) | 9.07 (2.29) | .010 | 0.093 |
| ADRD knowledge | 20.76<br>(4.01) | 21.32<br>(4.16) | 20.48<br>(3.91) | <.001 | -0.130 |
| Satisfaction with non-peer support sources | 2.52 (0.70) | 2.38 (0.72) | 2.59 (0.68) | <.001 | 0.184 |

The largest between-group differences were observed for belief in the value of online peer support and eHealth literacy, suggesting that recent users were not only more favorable toward online peer support but also more confident in locating and using health information online. Differences in caregiver stress, caregiving competence, and non-peer support satisfaction were statistically significant but smaller in magnitude. Although ADRD knowledge also differed significantly between groups, this difference was modest and in the opposite direction, with non-users showing somewhat higher knowledge scores.

### 3.4 Factors Associated with Online Peer Support Use

To address H2, we fitted ANFHSU-guided nested logistic regression models to examine factors associated with online peer support use.

Figure 2 summarizes discrimination across the five nested models predicting online peer support use in the past three months. Discrimination improved as domains were added sequentially, with ROC AUC increasing from 0.698 in the predisposing-only model (M1; n=1,113) to 0.744 after adding beliefs about the value of online peer support (M2; n=1,113). Additional gains were observed after incorporating enabling resources (M3; ROC AUC=0.754; n=1,110) and need-related factors (M4; ROC AUC=0.776; n=1,110). The full model including additional technology and contextual measures (M5; n=1,108) achieved the highest discrimination (ROC AUC=0.806). Sample size decreased slightly across models due to complete-case estimation. Consistent with this pattern, model fit improved across the nested sequence, with lower AIC and higher McFadden’s pseudo-R² in later models, and the full model (M5) showing the best overall fit.

**Figure 2.**
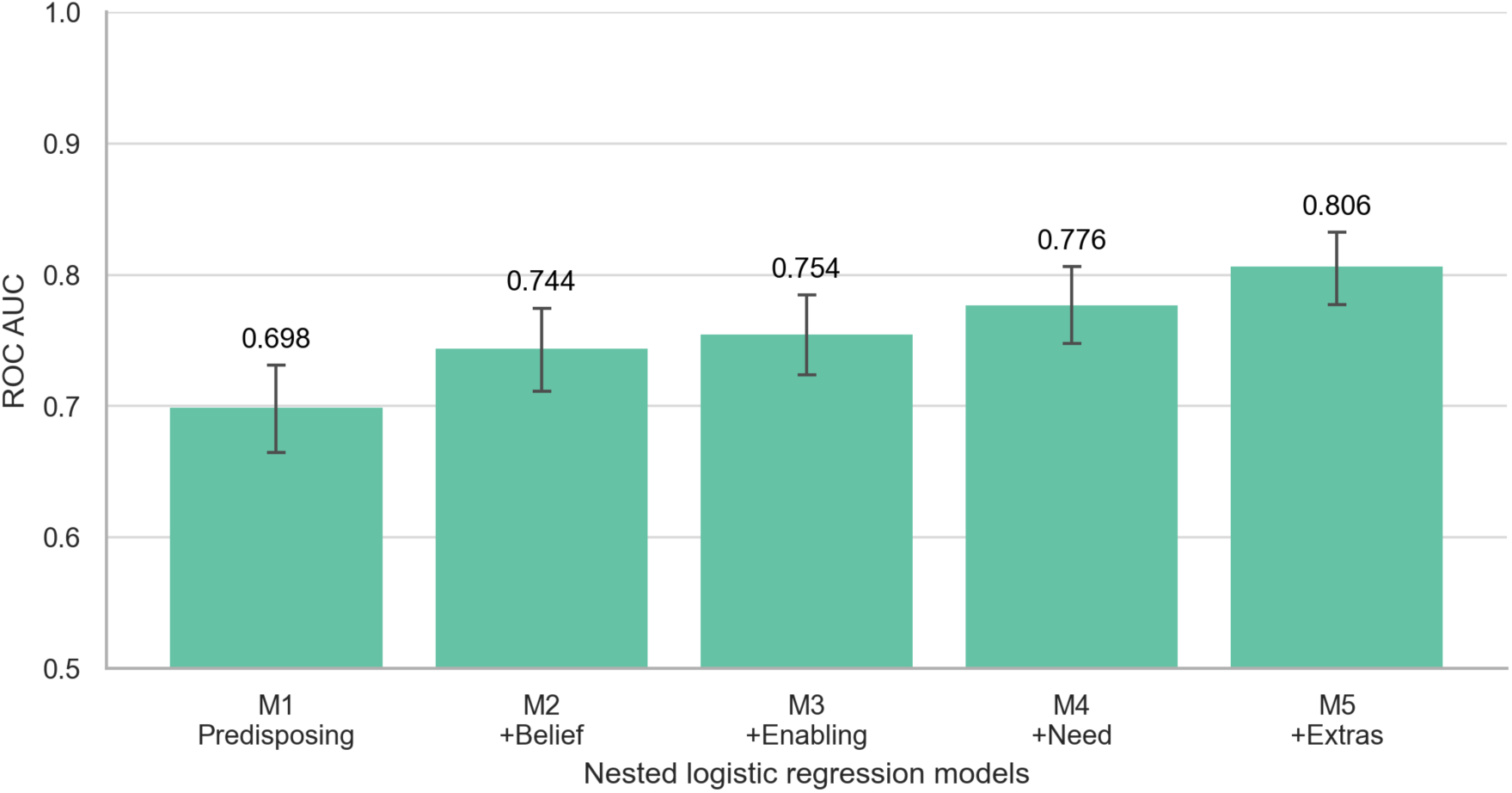
Discrimination across ANFHSU-guided nested logistic regression models predicting online peer support use in the past 3 months. Bars show ROC AUC for each model, and error bars show 95% bootstrap confidence intervals.

In the fully adjusted model (M5), seven factors were independently associated with online peer support use. Higher perceived usefulness was associated with higher odds of use (OR=1.04 per 1-point increase; 95% CI 1.01-1.08; p=.011). Enabling measures were also associated with use, including higher eHealth literacy (OR=1.06 per 1-point increase; 95% CI 1.02-1.10; p<.001) and greater willingness to use an experience-search tool (OR=1.68 per 1-point increase; 95% CI 1.39-2.04; p<.001). Higher caregiver stress was associated with increased odds of use (OR=1.03 per 1-point increase; 95% CI 1.01-1.05; p=.003). Several contextual factors were also associated with use: non-Hispanic/Latino caregivers had higher odds than Hispanic/Latino caregivers (OR=1.98; 95% CI 1.24-3.15; p=.004), part-time employed caregivers had higher odds compared with the reference employment group (OR=2.07; 95% CI 1.05-4.07; p=.035), and caregivers living in the same neighborhood as the care recipient had higher odds of use (OR=1.76; 95% CI 1.19-2.58; p=.004).

### 3.5 Nonuse Mechanisms: Belief-Behavior Gap and Reported Barriers

To address H3, we examined the belief-behavior gap among non-users by identifying a high-belief subgroup, comparing their profiles with other non-users, summarizing reported barriers, and modeling intention to use online peer support in the future. Figure 3 shows the distribution of perceived value (belief score) for online peer support among caregivers who did and did not report online peer support use in the past three months. Belief scores were higher among users, but the distributions overlapped substantially (Mann-Whitney U, P<.001; effect size=0.343). Among non-users (n=373), 108/373 (28.9%) met the Q75 high-belief threshold retained for detailed reporting, despite reporting no recent online peer support use.

**Figure 3.**
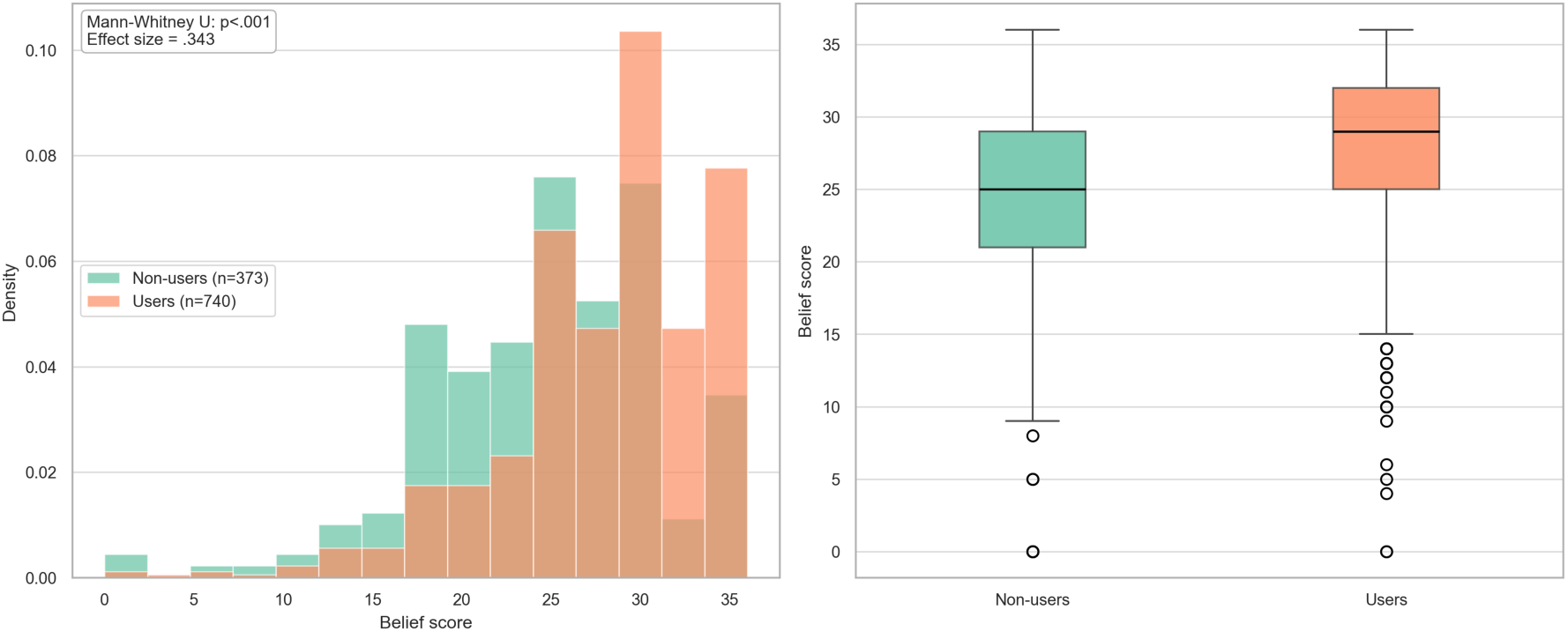
Distribution of perceived value (belief score) for online peer support among caregivers who did and did not report online peer support use in the past three months.

Compared with other non-users, high-belief non-users reported higher eHealth literacy (mean 28.4 vs 25.3; P<.001; Cohen’s d=0.57), greater willingness to use a caregiving-education tool (mean 4.4 vs 3.6; P<.001; d=0.83), greater willingness to use an experience-search tool (mean 4.0 vs 3.4; P<.001; d=0.61), higher caregiving competence (mean 9.5 vs 8.3; P<.001; d=0.47), and higher satisfaction with existing support (mean 2.7 vs 2.3; P<.001; d=0.57). Stress levels did not differ between groups (P=.43).

Open-ended responses from high-belief non-users suggested that nonuse often reflected friction in access and trust rather than rejection of online peer support itself. Several respondents described wanting a more efficient and credible pathway into online communities, for example requesting “a verified list of organizations to use” or “a shortlist of associations or online discussions” that could help them decide where to start. Others framed nonuse as a timing or workflow problem rather than a lack of interest, noting that “it would save me time second guessing the situation” or that they would use such support “at the beginning of the disease” or under more demanding caregiving circumstances. These responses suggest that some caregivers who value online peer support still do not engage because current options feel difficult to find, difficult to evaluate, or not sufficiently tailored to immediate caregiving needs.

Among all non-users, the most commonly reported reasons for not using online peer support in the past three months were already having sufficient offline support (118/373, 31.6%), lack of time to search posts or communicate online (102/373, 27.3%), disliking online communities (79/373, 21.2%), recently learning about online communities without a chance to join (74/373, 19.8%), and security or trust concerns (67/373, 18.0%). Additional barriers included not knowing where to find online communities (59/373, 15.8%), concerns that information in online communities may be misleading (57/373, 15.3%), and difficulty searching for needed information within known communities (32/373, 8.6%).

Among non-users, we additionally modeled intention to use online peer support in the next three months as a secondary outcome. Of the 373 non-users, 161 (43.2%) reported intending to visit online communities in the next three months. The intention model showed strong discrimination (ROC AUC=0.882), with McFadden’s R²=0.381 and AIC=419.9. Greater willingness to use a caregiving-education tool and greater willingness to use an experience-search tool were the strongest positive correlates of intention, whereas older age and caring for a person in late-stage dementia were associated with lower odds of intending future use.

## 4 Discussion

### 4.1 Principal Findings

This study provides an updated, ANFHSU-guided account of how informal caregivers of people living with ADRD use online peer support and what differentiates recent users from non-users. First, online peer support use was common, but engagement was heterogeneous. Caregivers reported using a fragmented set of platforms, with a long tail of less frequently named venues rather than concentration in only a few dominant sites. Engagement within these spaces also varied. More than half of recent users primarily read rather than post, indicating that online peer support often functions as a low-visibility, on-demand informational and emotional resource rather than a uniformly interactive community experience.

Second, online peer support use was associated with a combination of predisposing, enabling, and need-related factors. In the nested modeling sequence, model discrimination improved as additional domains were added, with a particularly notable gain after belief measures were introduced. In the fully adjusted model, higher belief in the value of online peer support remained independently associated with use, along with higher eHealth literacy, greater willingness to use an experience-search tool, and higher caregiving stress. The selected situational characteristics also correlate with service utilization. The use of services not only reflects caregiving needs but also embodies the caregiver’s perception of the value of online peer support, their ability to navigate digital resources, and the relevant practical conditions that enable their participation.

Third, we observed a meaningful belief-behavior gap. Belief scores were higher among users, but the distributions overlapped substantially, and a sizable subgroup of non-users still reported high perceived value despite no recent engagement. Compared with other non-users, these high-belief non-users showed stronger digital readiness and greater willingness to use digital support tools, including an experience-search tool and a caregiving-education tool, suggesting that nonuse cannot be explained solely by low capability or lack of interest. At the same time, non-users overall commonly reported barriers such as having already received sufficient offline support, limited time, limited opportunity to join despite awareness, and security or trust concerns. Open-ended responses further suggested that some caregivers wanted a more efficient, credible, and easier-to-navigate pathway into online communities. The findings indicate that favorable beliefs alone are insufficient to drive uptake when access, workflow, and trust-related frictions remain unresolved.

Finally, non-use was not always a stable or categorical position. A substantial proportion of non-users reported intending to use online peer support in the near future, and intention was more common among caregivers with greater willingness to use experience-search and caregiving-education tools. For many caregivers, nonuse appeared to reflect contingent barriers rather than rejection of online peer support itself. Future interventions should focus not only on persuading caregivers that online peer support is valuable, but also on reducing the effort required to find relevant communities, strengthening credibility and trust cues, and supporting passive participation as a legitimate and potentially beneficial form of engagement. Design and moderation strategies should also account for potential downsides of peer support, including misleading information, unconstructive or emotionally taxing exchanges, and advice that may be difficult to integrate with formal care.

### 4.2 Belief-Behavior Gap Analysis: Actual Barriers and Theories of Behavior

This study indicates a meaningful belief-behavior gap among informal caregivers of persons living with ADRD. Among 373 non-users, 28.90% met the high-belief threshold for online peer support effectiveness but did not use peer support during the study period. This pattern aligns with the Theory of Planned Behavior [36], which indicates that positive attitudes may still fail to produce action when perceived behavioral control is constrained by routine demands and contextual barriers. Within Andersen and Newman’s behavioral model [37], high-belief nonuse can be interpreted as a mismatch between predisposing factors, such as favorable beliefs and digital readiness, and enabling conditions, such as available time, trust, and practical access. Consistent with this interpretation, high-belief non-users reported higher eHealth literacy and greater willingness to use experience-search and caregiving-education tools than other non-users, suggesting that nonuse was not primarily explained by low digital capability alone. At the same time, commonly reported barriers among non-users included limited time, sufficient offline support, limited opportunity to join, and security or trust concerns, indicating that favorable beliefs may still fail to produce uptake when participation remains effortful or uncertain.

Social capital theory [38] helps explain why some caregivers who recognize the value of online peer support may still treat it as optional rather than necessary. Among high-belief non-users, 34 of 108 (31.5%) reported already having sufficient offline support, suggesting that existing bonding support may reduce the marginal value of additional online participation. This interpretation is consistent with network theory on weak ties [39], in which individuals may value informational breadth without seeking the relational investment implied by active participation. In practical terms, high-belief non-users may still recognize usefulness, but they may utilize online resources primarily as an “as-needed” reference rather than as a setting for ongoing reciprocity.

Trust and credibility concerns require an additional sociological explanation beyond “privacy preference.” Goffman’s stigma and impression-management framework [40] indicates that public participation carries identity risk, particularly when caregivers anticipate judgment about competence, family dynamics, or emotional control. This framing is consistent with our reported security and credibility concerns, because the perceived risk is not only technical (data privacy), but also social (unwanted visibility, misinterpretation, and loss of face). In parallel, research on medical information overload and information avoidance indicates that high-volume, low-certainty information environments can increase cognitive burden and promote disengagement even among highly motivated individuals [41–44]. Consistent with the Health Belief Model [45], caregivers may require cues to action, such as acute symptom escalation, care transitions, or clinician guidance, to cross a higher participation threshold when time scarcity and credibility uncertainty remain dominant.

### 4.3 Passive Participation: Lurking as Structured Engagement

The finding that 54.3% of users primarily read rather than post indicates that Passive Participation [46] is a central mode of engagement in caregiver peer support. This pattern is consistent with longstanding work on participation inequality in online communities [47], and it suggests that posting behavior is not an appropriate sole indicator of benefit or involvement. Research on lurkers indicates that reading is often purposeful, including information seeking, risk management, and norm learning, rather than simple disengagement [48–50]. In the context of ADRD caregiving, this interpretation is especially plausible because caregivers experience high time scarcity and may prioritize efficient “just-in-time” informational support [51].

Social learning theory [52] directly explains why reading might be an expected first-stage behavior. Lave and Wenger’s Legitimate Peripheral Participation suggests that newcomers typically begin at the periphery, observing normative, linguistic, and credibility cues before gradually deepening their engagement [53]. Applied to caregiver peer support, reading can be interpreted as a low-risk strategy for assessing the relevance of community experiences, the credibility of advice, and the controllability of emotional tone. This mechanism also aligns with Anderson and Newman’s model, as even caregivers with strong perceived needs may encounter constraints such as time constraints and uncertainty about information disclosure norms, making posting difficult.

In addition, work on the *economies* of online cooperation [54] indicates that contribution depends on perceived value, expected reciprocity, and the perceived costs of producing a high-quality post. In caregiver settings, those costs include not only time, but also the emotional labor of narrating distressing experiences and the uncertainty of how others will respond. Lurking can function as identity protection, allowing caregivers to access information while minimizing visibility and reducing the risk of judgment or unwanted exposure [55]. This interpretation is consistent with the broader finding that trust and security concerns remain salient barriers [56].

If reading is a legitimate and common avenue for engagement, digital interventions should not treat lurkers as unsuccessful participants. Platforms may instead support gradual participation through clearer onboarding, structured prompts, question templates, search and navigation support, and credibility cues. These considerations are consistent with prior dementia caregiver technology research emphasizing flexibility in delivery, information quality, ease of use, and supportive connectedness [11,35]. Platform structure and moderation may also shape engagement, as more organized, condition-specific communities may provide clearer norms and credibility cues than broader social platforms [51]. Intervention evaluation should likewise use a broader engagement model that treats reading frequency, bookmarks or saves, and content navigation as meaningful usage metrics.

### 4.4 Limitations and Future Work

This study has several limitations. First, the cross-sectional design limits inference about the temporal ordering of beliefs, enabling resources, caregiving-related need, and online peer support use. Future longitudinal studies are needed to clarify how these factors shape uptake and sustained engagement over time.

Second, the survey relied on a national, nonprobability Qualtrics panel sample rather than a probability-based sample. Although this approach enabled efficient recruitment of a large national caregiver sample, it may not represent all ADRD caregivers, particularly those with limited internet access, lower digital literacy, or less willingness to participate in online panels. Findings should therefore be interpreted as observational patterns within a web-recruited caregiver sample rather than population estimates. Future studies should combine broader community and clinical recruitment strategies with methods that better assess equity-relevant barriers.

Third, key measures were self-reported and may be subject to recall error or social desirability bias, particularly for online behaviors and intended future use. Future work could strengthen measurement by incorporating complementary approaches such as behavioral tasks, passive usage metrics when feasible, or platform-level analytics.

## 5 Conclusions

Online peer support was commonly used by informal caregivers of people living with ADRD, but engagement was heterogeneous, distributed across a fragmented platform ecosystem, and often characterized by passive participation. Within an ANFHSU framework, recent use was associated with perceived value, digital readiness, and caregiving-related need, suggesting that uptake depends not only on motivation but also on whether online support is accessible, credible, and feasible within the constraints of caregiving. A meaningful belief-behavior gap was also evident: some caregivers who viewed online peer support as valuable still did not use it, reflecting barriers related to time, trust, discoverability, and existing offline support. Efforts to support ADRD caregivers should therefore reduce friction in finding relevant and credible communities, strengthen trust and usability, and design for passive as well as active forms of engagement.

## Data Availability

The data are not publicly available due to privacy and ethical considerations. Deidentified data may be available from the corresponding author upon reasonable request and subject to applicable institutional and ethical requirements.

